# Influence of heterogeneous age-group contact patterns on critical vaccination rates for herd immunity to SARS-CoV-2

**DOI:** 10.1101/2021.11.03.21265897

**Authors:** Joan Saldaña, Caterina Scoglio

## Abstract

Currently, several western countries have more than half of their population fully vaccinated against COVID-19. At the same time, some of them are experiencing a fourth or even a fifth wave of cases, most of them concentrated in sectors of the populations whose vaccination coverage is lower than the average. So, the initial scenario of vaccine prioritization has given way to a new one where achieving herd immunity is the primary concern. Using an age-structured vaccination model with waning immunity, we show that, under a limited supply of vaccines, a vaccination strategy based on minimizing the basic reproduction number allows for the deployment of a number of vaccine doses lower than the one required for maximizing the vaccination coverage. Such minimization is achieved by giving greater protection to those age groups that, for a given social contact pattern, have smaller fractions of susceptible individuals at the endemic equilibrium without vaccination, that is, to those groups that are more vulnerable to infection.

## Introduction

The pandemic of the severe acute respiratory syndrome coronavirus 2 (SARS-CoV-2) is still imposing incredible pressure on many countries’ healthcare and economic systems.

Nations in America, Europe, Asia, and Africa have faced large numbers of deaths due to COVID 19, and a continued crisis situation. The only good news in this dark situation is that vaccines are becoming available from different companies all over the world. Some countries are currently evaluating the efficacy and effectiveness of developed vaccines, while some other countries have already started their vaccination campaign. In particular, as of November 2021, more than 75% of the people in countries like Canada, United Kingdom, and France have received at least one dose of the vaccination, while no more than 35% of people in Bangladesh, Guinea, and Armenia have received one dose at least^1^. These examples give an idea of the heterogeneous situation of the vaccination campaign in the world. Additionally, current studies suggest a decay of neutralizing antibody responses in convalescent patients^2^, as well as a decline in the effectiveness of mRNA COVID-19 vaccines^3^. Therefore, vaccines probably provide a short-lived immunity. For instance, comparing the rate of decay of the antibody responses following infections by human coronavirus (hCoV) and by SARS-CoV-2, it has been suggested that individuals may become susceptible to reinfection within 12-18 months after a previous infection^2^. Similarly, a recent comparative evolutionary analysis of coronavirus relatives of SRAS-CoV-2 reveals that, under endemic conditions, reinfection by SARS-CoV-2 would likely occur between 3 months and 5.1 years after peak antibody response, with a median of 16 months^4^.

While during the ongoing pandemic most countries agree to vaccinate first public health personnel and people in long term care facilities, the limited availability of vaccines and the logistic complexities are still posing big questions on when and how the vaccination campaign will be completed. Initially, some countries assessed reaching herd immunity at around 70% of the population vaccinated^5^. With time, several hurdles upon this achievement are becoming evident^6^. These difficulties in obtaining herd immunity may further discouraging people from attaining it.^5^.

Countries are now developing immunization plans to face the challenge of distributing millions of vaccines, some of which require very special maintenance conditions. These plans include the definition of priority schemes to start the distribution process and, since it is very likely that not all people will be vaccinated for different reasons, understanding how vaccine distribution among population age segments impacts the obtained herd immunity is of primary importance.

A vaccination strategy can be developed to obtain different goals such as minimizing deaths, minimizing number of cases, minimizing severe cases requiring hospitalization, etc.^7^. For instance, in a very recent paper^8^, the authors consider five different vaccine prioritization strategies. Among their findings, it was shown that giving priority to adults ages 20-40 years minimizes the cumulative incidence, while mortality is minimized by giving priority to adults ages 60 and more. Another very recent article^9^ investigates two criteria for vaccination priority based on age: lives saved and years of future life saved. While in general these two criteria can be in conflict, in the case of COVID-19, vaccinating the oldest first saves the most lives and simultaneously also maximizes years of remaining life expectancy.

In another approach to vaccination^10^, the authors have investigated how much vaccine is required by any given country, year by year, to create herd immunity to block SARS-CoV-2 transmission, assuming immunity is short lived (waning immunity). To answer this question, a simple model is developed showing the percentage of the population in the first year of an epidemic that must be vaccinated and the percentage that must be vaccinated once the system reaches equilibrium after a few years. Results show that in year 1 a much larger fraction of the population needs to be vaccinated, being most of the population susceptible, compared with the population fraction to be vaccinated in subsequent years, to create effective herd immunity.

The study of vaccination strategies to achieve herd immunity has been considered for several infectious diseases and for many years^11–13^. In particular, vaccination schemes aiming to reduce *R*_0_ below its threshold value 1 are called *preventive* because, once they have been launched, epidemic outbreaks are not possible under occasional introduction of new cases in the population. For stochastic SIR epidemics with permanent immunity, preventive strategies have been considered, for instance, in^14^. However, due to the lack of vaccines at the beginning of the COVID-19 pandemic, some papers analysed the possibility of reaching disease-induced herd immunity in age-structured models. In a recent paper^15^, an SEIR model has been developed to assess the feasibility of suppressing the virus transmission or, alternatively, of achieving herd immunity, by applying social distancing to differing age groups and self-isolation by symptomatic infectious individuals. The model revealed that obtaining herd immunity without exceeding hospital capacity was not a practical objective because, without a vaccination program, social distancing needed to be maintained for an extended period and adapted over time in a “precise yet unfeasible way”. Almost at the same time, a second paper^16^ has adopted a similar approach to study the level of immunity that can be achieved with non-pharmaceutical interventions. Its aim was to relate the severity of preventive measures imposed at the beginning of the pandemic with the size of outbreaks appearing after these measures were lifted. In this setting, it is shown that, when the age structure of the focal population is considered, herd immunity can be reached at around 43% instead of the traditional value of 60% that appears for a basic reproduction number *R*_0_ = 2.5 under a homogeneous mixing of the population. In both papers^15,16^, social distancing interventions are modeled by reducing the mean number of contacts in the original contact matrix.

In this paper, we deal with the question of the challenges associated to creating herd immunity to SARS-CoV-2 infection by means of preventive vaccination strategies with waning immunity that take into account the contact rates among age segments. In particular, short-lived immunity implies that continuous vaccination campaigns are needed to preserve the herd immunity. Therefore, we adopt the assumption of reaching a disease-free equilibrium (DFE) where susceptible and vaccinated individuals are only present^10^. Then, using an age-structured Susceptible-Infected-Recovered-Vaccinated model, we firstly derive the expression for the vaccination rates that lead to the maximum vaccination coverage at this equilibrium for a given supply of vaccines per unit time (the total vaccination rate). Next, if 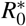 denotes the basic reproduction number at the DFE with vaccinated individuals, we compute two different sets of per age-group vaccination rates: 1) the set that minimizes 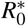 with the constraint that the total vaccination rate is the same as the critical rate under uniform vaccination, and 2) the set at which the minimum 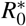 equals 0.996 when a suitable (and lower) total vaccination rate is assumed.

For the limited supply of vaccine given by the critical vaccination rate under a homogeneous mixing, we found that the value of 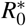 obtained by maximizing the vaccination coverage is always larger than the minimum of 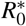 attainable under the same constraint on the total vaccination rate. The latter then defines the optimal allocation of vaccines among age groups under the given supply. On the other hand, since this minimum 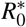 will be clearly less than 1, the vaccination rates of the second set 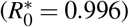 will be smaller than those of the first set, thus achieving the herd immunity at a lower supply of vaccine. We have also verified that these results hold when considering (1) a high but not complete success rate for vaccines, and (2) a different duration of immunity for each age group, in accordance with observations of the age-related decline of the immune system that weakens the ability to mount effective responses to vaccines. This decline is often referred to as “immunosenescence”^17,18^.

To our knowledge, this is the first study on the combined impact of age-group contact patterns and short-lived vaccination immunization on the optimal allocation of vaccines among age groups. We are able to quantify the importance of specific contact patterns in different countries through the reduction of 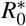 at the optimal strategy that results when vaccination by age group is used instead of the one based on a uniform distribution of vaccines^10^. In contrast to other preventive vaccination strategies, like the one aiming to maximize the vaccination coverage under a given supply of vaccines, minimizing 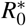 with respect to the set of per capita vaccination rates leads to preferential targeting for the most vulnerable age groups, that is, those with higher contact rates. Although the importance of targeting highly connected individuals to reduce the virus transmission is well known in epidemiology^19^ and, in particular, in the so-called contact network epidemiology^20^, this is the first time that the criterion of minimizing 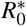 is used to find an optimal vaccine allocation among age groups. Even though we are aware that the numerical results are obtained on the base of measured and consequently noisy contact patterns, our mathematical model reveals the critical role played by the age-based contact patterns in efficiently administering vaccines and can be useful in encouraging the population to see a possible end of the pandemic by vaccination.

### The model

In this paper we consider a deterministic epidemic model with continuous vaccination where individuals are classified in three age groups: youngsters, adults, and the elderly (*i* = 1, 2, 3 respectively). Within each age class, individuals are classified according to their disease status: susceptible, infectious, recovered, and vaccinated. It is assumed a loss of immunity in recovered and vaccinated individuals at rates *δ*_*i*_ and 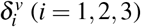, respectively. Moreover, as mentioned at the Introduction, the probability that the vaccine successfully protects against infection is assumed to be age-dependent and it is denoted by *p*_*i*_ (*i* = 1, 2, 3).

The number of infections in age group *i* caused by individuals of age group *j* is *β c*_*i j*_*S*_*i*_*I*_*j*_*/N*_*j*_, where *S*_*i*_ is the number of susceptible individuals in age group *i, I*_*j*_*/N*_*j*_ is the fraction of infected individuals in age group *j, β* is the transmission probability through an infectious contact, and *C* = (*c*_*i j*_) is the social contact matrix. *C* gives the (mean) number of contacts per unit time between an individual of age group *i* and individuals of age group *j*, and is the central ingredient of the model since it reflects how individuals mix with each other in different countries.

To model the limited capacity of a public health system or situations of vaccine shortage as those occurring in low-income countries where fewer than 3% of people has been fully vaccinated against COVID-19 as of November 2021^1^, a fixed number *w* of individuals is assumed to be vaccinated per unit of time and, moreover, we will assume an age-dependent targeting of vaccination^8, 12, 21^. So, if *w*_*i*_ denotes the number of vaccines assigned to age group *i* per unit of time (vaccination rate of age group *i*), then *w*_*i*_ *S*_*i*_*/N*_*i*_ is the number of susceptible individuals in age group *i* vaccinated per unit of time when vaccination is made regardless of disease status^22^. In particular, if we do not consider age, then under a uniformly random vaccination of a population it follows that *w*_*i*_ = *w f*_*i*_ with *f*_*i*_ = *N*_*i*_*/N*, i.e., *w*_*i*_ is proportional to the fraction of the population in age group *i*, with 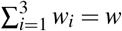, the total vaccination rate. In such a case, the vaccination term in the equation for the susceptible individuals in age group *i* is given by *w*_*i*_ *S*_*i*_*/N*_*i*_ = *w f*_*i*_ *S*_*i*_*/N*_*i*_ = *w S*_*i*_*/N*. So, any strategy that departs from this uniform vaccination will be given by a vector (*w*_1_, *w*_2_, *w*_3_) of vaccination rates satisfying 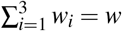.

According to these hypotheses, assuming an arbitrary vaccination strategy (*w*_1_, *w*_2_, *w*_3_), and ignoring the demographics of aging, birth, and death given the short timescale of an epidemic, the equations governing its dynamics are

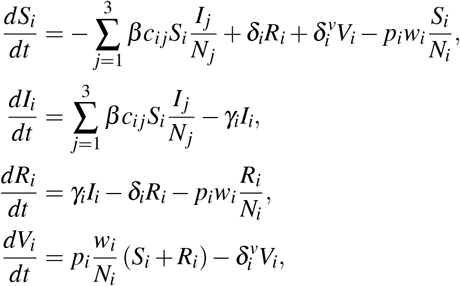

with *S*_*i*_ + *I*_*i*_ + *R*_*i*_ + *V*_*i*_ = *N*_*i*_, *i* = 1, 2, 3, and 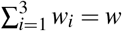.

Writing the system in terms of fractions (*s*_*i*_ = *S*_*i*_*/N*_*i*_, *y*_*i*_ = *I*_*i*_*/N*_*i*_, *r*_*i*_ = *R*_*i*_*/N*_*i*_, and *v*_*i*_ = *V*_*i*_*/N*_*i*_) and neglecting the last equation because it is redundant, we have

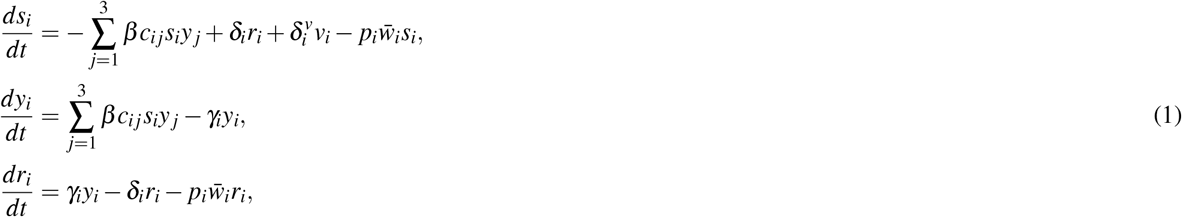

with *s*_*i*_ + *y*_*i*_ + *r*_*i*_ + *v*_*i*_ = 1, and 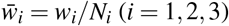 being the per capita vaccination rate of age group *i*. Note that, from the constraint 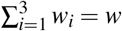 and the definition of 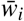, it follows that 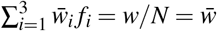, the mean per capita vaccination rate in the population.

The previous relationship among the per capita vaccination rates of each age group implies that, if the population fraction of an age group *i* is lower than the number 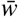 of available vaccines per person and per unit of time 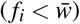, then 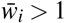 is required when vaccines are mainly targeted at this age group (i.e., when 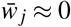 for *j* ≠ *i*). Roughly speaking, 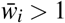 corresponds to situations in which there are more available vaccines per unit of time than people to be vaccinated in the *i*-age group (remember we are assuming a constant vaccination rate *w*).

#### The disease-free equilibrium and vaccination strategies

The disease-free equilibrium (DFE) of system (1) is 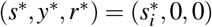 where

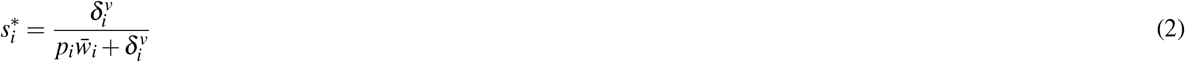

with 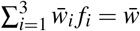. Therefore, at this equilibrium, only susceptible and vaccinated individuals are present with 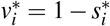 (*i* = 1,2,3).

The basic reproduction number at the DFE, here denoted by 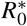 to distinguish it from the one for the model without vaccination, is the largest eigenvalue of the next-generation matrix^23^

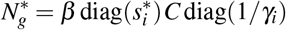

where 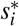 is given by (2), *C* is the social contact matrix, and 1*/γ*_*i*_ is the mean infectious period of infected individuals of age group *i*. Moreover, since the per capita vaccination rates 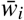 satisfy 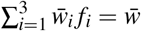 (limited supply of vaccines), 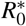 can be considered a function of 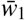 and 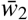 only, that is, 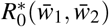.

From (2) we can compute the condition on 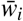 for having a maximum vaccination coverage of the population at the DFE, which is equivalent to minimize the fraction of susceptible population at this equilibrium, 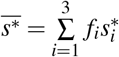. Precisely, the condition grad 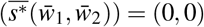 and the positivity of the rates amount to

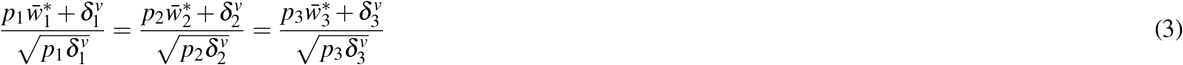

with 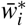 satisfying 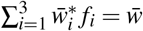. So, from (3) one easily obtains an explicit expression for the vaccinations rates 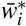 leading to the maximum vaccination coverage under the constraint of having a given amount of vaccine per unit of time, which turns out to be a global maximum. In particular, it follows that, if the rate of immunity loss 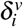 is the same for the vaccinated individuals of all the age groups and the probability of being protected after vaccination *p*_*i*_ is also the same across age groups, then the vaccination rates that guarantee the maximum fraction of vaccinated population are 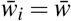, i.e., they correspond to a uniformly random vaccination of the population, irrespective of age.

By definition, under such a vaccination strategy, the probability of being vaccinated per unit of time is the same for any individual regardless of age. In this case, the differential equation governing the dynamics of the susceptible individuals of age group *i* is

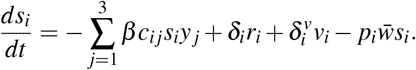

Using 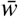 as a tuning parameter, we compute the basic reproduction number 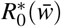, and find the critical per capita vaccination rate 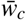 defined by 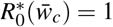. As an example, Fig. 1 shows the behaviour 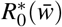 with the data set from Italy which leads to 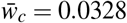.

**Figure 1.**
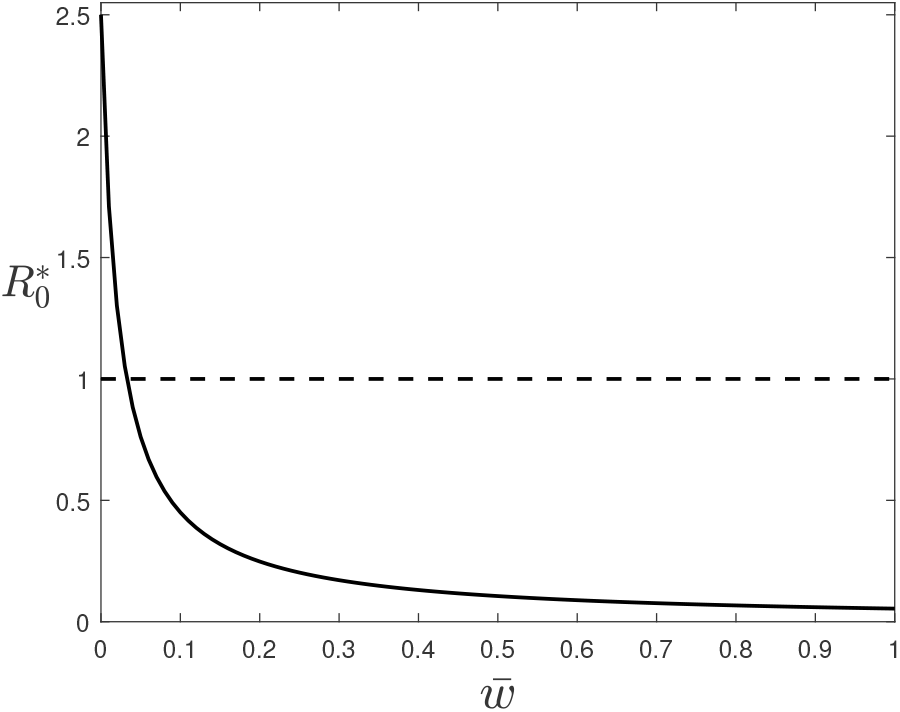
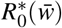 for system (1) at the disease-free equilibrium given by (2). The critical vaccination rate is given by the intersection of 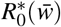 with the dashed line 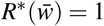. Parameters: 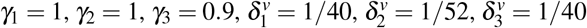, *p*_1_ = *p*_2_ = *p*_3_ = 1, and *β* is scaled such that *R*_0_ = 2.5 for the data set from Italy in the absence of vaccinated individuals 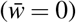, see Results and Discussion for details.

When the rates 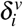 of immunity loss among groups differ from each other, the maximum vaccination coverage at the DFE will be attained for values of the per capita vaccination rates 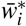 that do not correspond to a uniformly random vaccination of the population. These 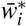 are optimal in the sense that they maximize the vaccination coverage; however, they do not guarantee the minimum value of 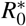 at the DFE. This fact, indeed, can be used to define an alternative criterion for an optimal vaccination strategy, namely, the one than leads to the lowest value of 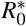 at the DFE. Under such a strategy, we control the disease by targeting age groups according to their potential contribution to an epidemic outbreak. So, we use the same amount of vaccines per unit of time but, in comparison to the random vaccination, we are vaccinating more individuals from some age groups while other age groups are less vaccinated. This situation corresponds to what has been called an optimal but inequitable distribution of vaccine^7^.

## Methods

### Data

The Social Contact Data initiative (http://www.socialcontactdata.org) includes contact matrices for Belgium, Finland, Germany, Italy, Luxembourg, Netherlands, Poland and the UK from POLYMOD^24^, as well as data from studies on social mixing in other countries^25^. All data are available on Zenodo at https://doi.org/10.5281/zenodo.1215899, and can be retrieved within R using the socialmixr package. The SOcial Contact RATES (Socrates) data tool at http://www.socialcontactdata.org/socrates/^26^ enables quick and convenient retrieval of these social contact matrices. Using the Socrates data tool, we have selected six countries for our numerical evaluations: Belgium, Germany, Italy, the Netherlands, Peru, and Zimbabwe. All the European data-sets are from the POLYMOD project^24^. The contact matrix for Peru is from^27^ and the one for Zimbabwe is from^28^.

We have selected three age groups: the first group includes people of age 0 to 17 years, the second group includes people of age 18 to 59 years, and the third group includes people 60 years old and older. For each of these countries, the Socrates data tool provides a matrix containing the mean daily number of contacts an individual of age group *i* (row) has with individuals of age group *j* (column). The row sums of this matrix correspond to the total per capita contact rate of each age group. In all these matrices, contacts are reciprocal (see Supplementary Table S1 online for more details).

The four European countries have a similar population composition with 20-25% of people in the third age group (the elderly) and about 57% in the second age group (adults), which is in sharp contrast with the younger populations of Peru and Zimbabwe where the elderly only represents 8.9% and 4.4% of the population, respectively (see Supplementary Table S2 online). However, the contact patterns of these four European countries show clear and important differences. Two of them, Italy and the Netherlands, have very dissimilar per capita contact rates, with very high daily numbers of contacts among children and among adults, whereas Belgium and Germany have lower and less dissimilar per capita contact rates. On the other hand, in Zimbabwe, elderly people are the age group with the highest total per capita contact rate, whereas the same age group has the lowest total per capita contact rate in the other countries. In Peru, the total per capita daily number of contacts in the first age group almost doubles the total per capita daily number of contacts in the other two age groups.

### Parameters

The recovery rate and loss of immunity rate are assumed to be the same in the six countries in exam and, also, to be very similar among groups. The recovery rates are *γ*_1_ = *γ*_2_ = 1 and *γ*_3_ = 0.9. Therefore, since these rates are equal to 1 (or very close to it), we can consider that time is measured in units of the infectious period, which is about one week (8 to 10 days) for moderate cases of COVID-19^29^. The loss of immunity rates for recovered (*δ*_*i*_) and vaccinated 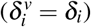 individuals are *δ*_1_ = *δ*_3_ *> δ*_2_ = 1*/*52. These values of 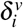 say that, on average, individuals between 18 and 60 years have one year (52 weeks) of immunity against the disease, while the length of this period is assumed to be shorter for individuals from the other two age groups (40 weeks; except for Supplementary Fig. S1 online where it is equal to 26 weeks). These shorter periods reflect the lower maturity of the immune system in the first age group^30^ and the immunosenescence in the elderly^18^.

For each country, the transmission rate *β* is obtained by imposing that *R*_0_ = 2.5 at the beginning of the epidemic when vaccinated individuals are not present. In this case, *R*_0_ is the largest eigenvalue of the next-generation matrix

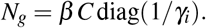

The original matrix *C* is given in terms of contacts per day. Since our unit of time is one week, we multiply the contact matrices in the Appendix by 7 to compute *β*. However, note that working with these re-scaled contact matrices only affects the value of *β* (which is divided by 7 when re-scaled contact matrices are used) but not the results we are presenting because the product *βc*_*i j*_ in the incidence term is invariant to the re-scaling of *C*.

## Results

### Without vaccination

Given that the disease parameters across the countries are assumed to be the same, we can assess the impact of the social contact patterns on the disease spread and, in particular, on the optimal vaccination strategy. Fig. 2 shows the evolution of the fraction of susceptible individuals during an epidemic without vaccination for the selected parameters. Although the mean fraction is always very similar (we are imposing the same value of *R*_0_ in all considered countries), we can see that the smallest (highest) fraction of susceptible individuals always corresponds to the age group with the highest (lowest) total per capita contact rate (see the last column of Supplementary Table S1 online). This figure also reveals that the similar contact patterns in Italy and the Netherlands lead to the same ordering of the fractions of susceptible individuals with respect to the mean fraction.

**Figure 2.**
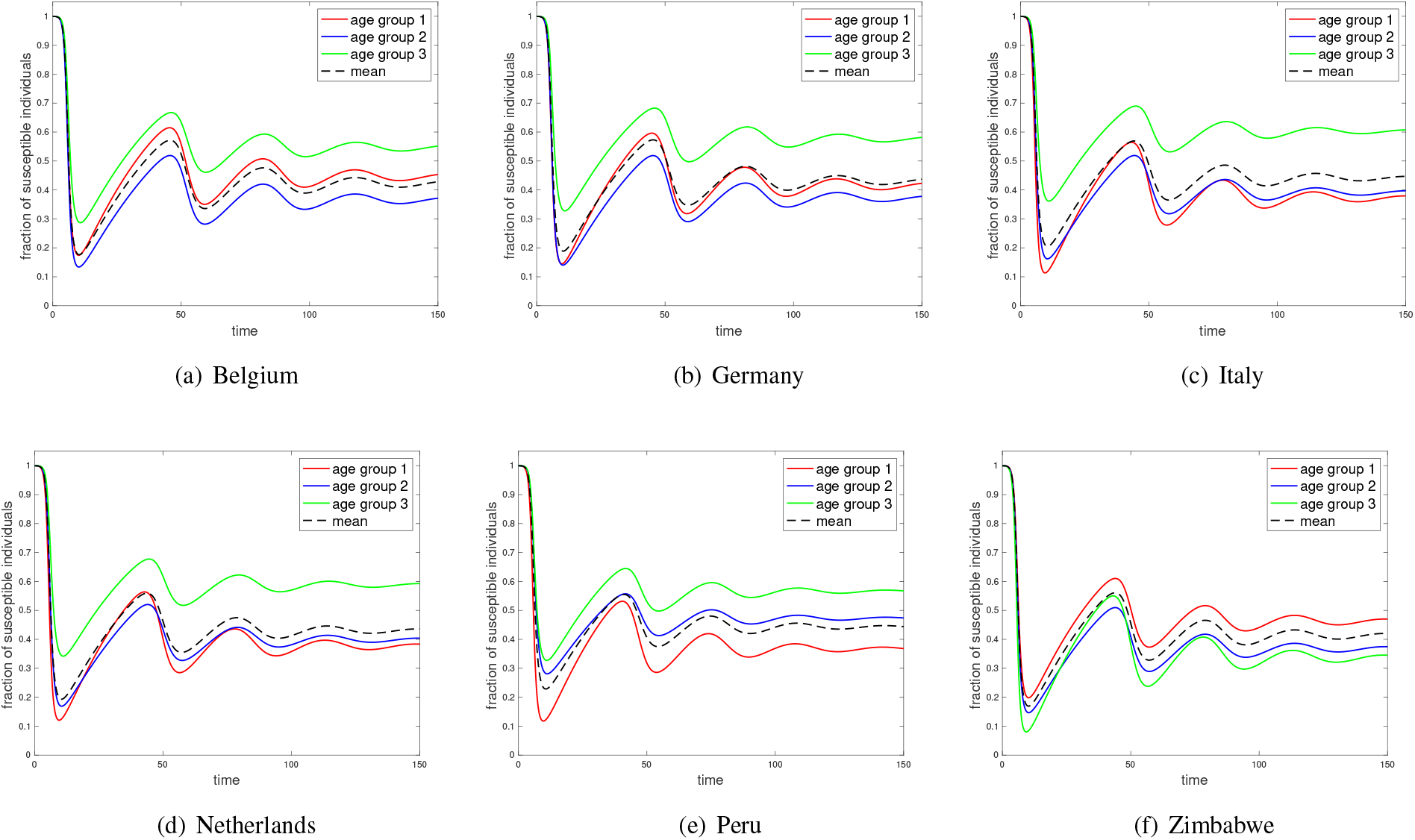
Examples of evolution of susceptible individuals during an epidemic given by (1) without vaccination with initial condition (*s*_*i*_(0), *y*_*i*_(0), *r*_*i*_(0)) = (0.9999, 0.0001, 0) for *i* = 1, 2, 3. Parameters: *γ*_1_ = 1, *γ*_2_ = 1, *γ*_3_ = 0.9, *δ*_1_ = 1*/*40, *δ*_2_ = 1*/*52, and *δ*_3_ = 1*/*40. For each country, *β* is scaled such that *R*_0_ = 2.5 for the corresponding data set in the absence of vaccinated individuals.

### Critical rates under uniformly random vaccination

For all the data sets, the critical per capita vaccination rate under the uniformly random vaccination 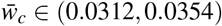 when perfect protection is assumed. The specific critical rate values and vaccination coverage for each country are reported in Table 1 under full protection of the vaccine (the two left columns), and probabilities *p*_1_ = 1, *p*_2_ = 0.95, and *p*_3_ = 0.9 of being protected after vaccination (the two right columns). In both scenarios, Peru is the country with the largest critical per capita vaccination rate (3.54% and 3.59%, respectively), which leads to the highest vaccination coverage of the population (62.03% and 61.48%, respectively). We can interpret these values of the coverage as the herd immunity level required for Peru under a uniformly random vaccination.

**Table 1.** Mean vaccination coverage (in %) given by (4) adopting the uniformly random vaccination strategy at the critical per capita vaccination rate with a 100% vaccine efficacy, 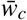 (in %), and with probabilities p = (1, 0.95, 0.9) of being successful, 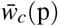 (in %). Parameters: 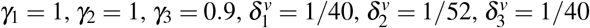. For each country, *β* is scaled such that *R*_0_ = 2.5 without vaccination.

| Data set | $\bar{w}_c$ | $V_{\bar{w}_c}$ | $\bar{w}_c(\mathbf{p})$ | $V_{\bar{w}_c}(\mathbf{p})$ |
| --- | --- | --- | --- | --- |
| Belgium | 3.1222 | 59.16 | 3.2678 | 58.99 |
| Germany | 3.1487 | 59.37 | 3.2836 | 59.04 |
| Italy | 3.2765 | 60.36 | 3.3823 | 59.77 |
| Netherlands | 3.3374 | 60.86 | 3.4246 | 60.27 |
| Peru | 3.5408 | 62.02 | 3.5898 | 61.47 |
| Zimbabwe | 3.2882 | 59.73 | 3.4161 | 59.98 |

Table 1 shows that, as expected, the critical vaccination rate increases when there is a fraction of people who are not completely protected after being vaccinated (*p*_*i*_ *<* 1 for *i* = 2, 3). It also shows a small decrease in the vaccination coverage for all the data sets except for the one from Zimbabwe. Recall that, under uniformly random vaccination, 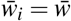 for *i* = 1, 2,3. Hence, the mean vaccination coverage at the DFE given by (2) with a critical per capita vaccination rate 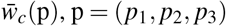, is given by

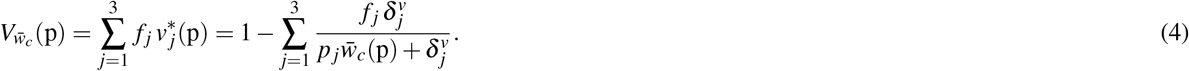

Note that 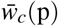 decreases when the probabilities *p*_*i*_ of being protected after vaccination increase. So, the dependence of 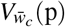 on *p*_*i*_ is through the products 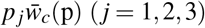. From the critical vaccination rates in this table and the corresponding values of *p*_*i*_, it follows that 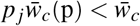 for *j* = 2,3. This means that the first age group is the only one with a higher percentage of vaccinated individuals at the DFE when *p*_2,3_ *<* 1. Zimbabwe, moreover, is the country with the highest fraction of people in this age group (49.1%, see Supplementary Table S2 online). These two facts explain why the mean vaccination coverage increases only in Zimbabwe when an imperfect protection of the vaccine is assumed for the second and third age groups.

### Vaccination strategies

In Fig. 3 we show the contour plots of 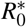 as a function of 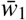 and 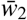 for the six data sets. In this figure, we assume that the mean vaccination rate 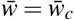 and 100% of vaccine efficacy. Because of the constraint 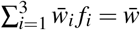, the interior boundary of each plot corresponds to 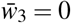, i.e., to straight line given by 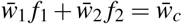.

**Figure 3.**
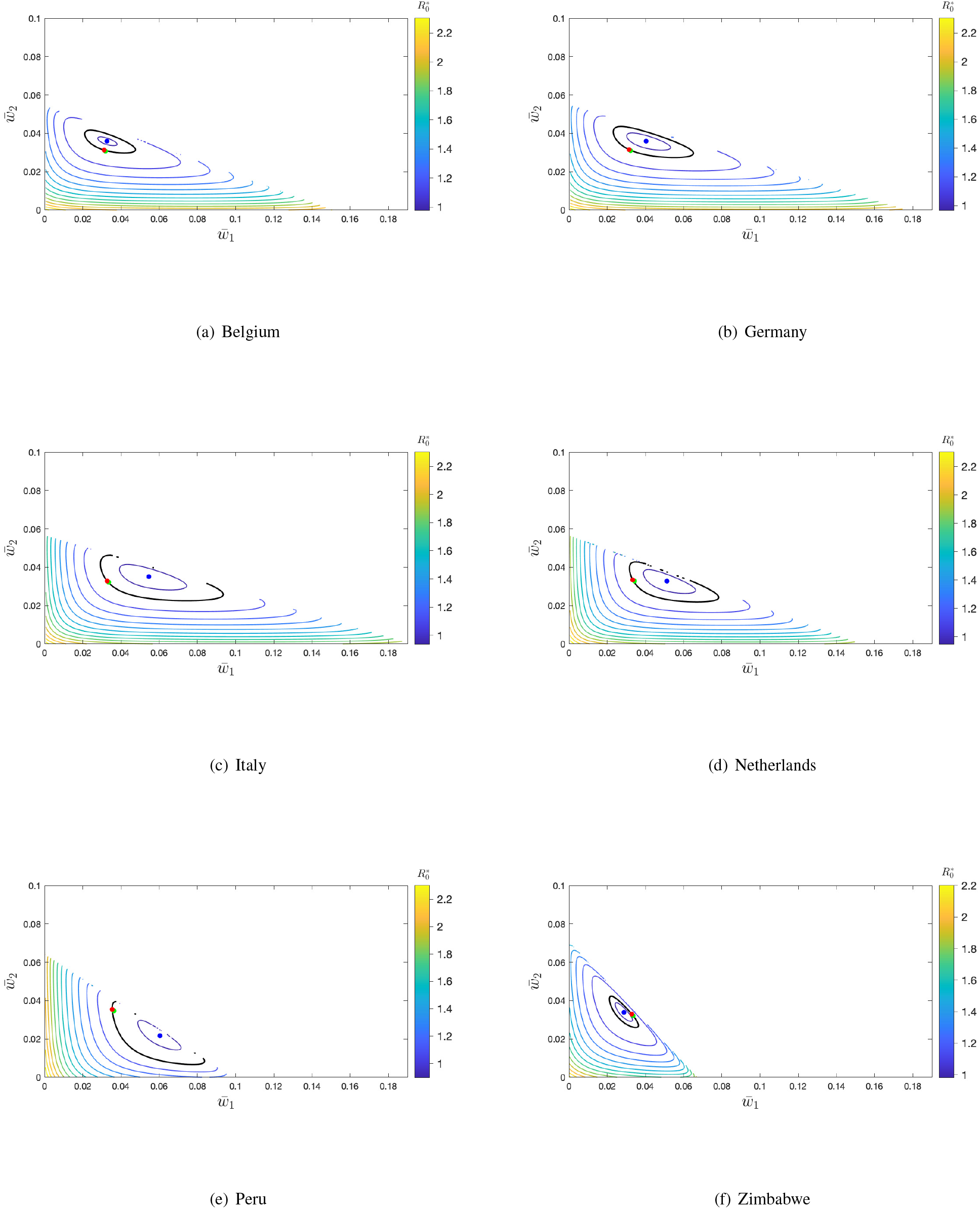
Contour levels of 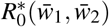 at the DFE of system (1) with *p*_*i*_ = 1 (*i* = 1, 2, 3). Black level curve corresponds to 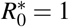. Blue point: Minimum of 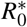. Red point: 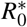 under uniformly random vaccination 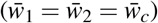. Green point: 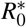 at the disease-free equilibrium with the maximum vaccination coverage. Parameters: 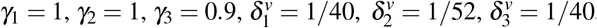, and 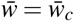 for each data set. For each country, *β* is scaled such that *R*_0_= 2. 5 in the absence of vaccinated individuals.

It is interesting to observe that the population fraction of the second age group (18 to 59 years), *f*_2_, varies narrowly from 0.46 (Zimbabwe) to 0.59 (Netherlands). This is why the range of values of 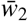 goes from 0 to 0.055-0.071 in all panels of Fig. 3 (its maximum value is 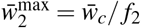). By contrast, the values of 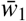 (and, so, those of 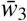) show a greater variability. The most extreme situation appears in the data from Zimbabwe with the highest population fraction in the first group (0-17 years) 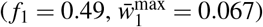, and the lowest fraction in the third group (60+ years) 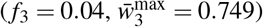.

With respect to the vaccination strategies shown in the panels of this figure, the (red) point 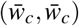 corresponds to the uniformly random vaccination with 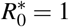 and, as expected, it is very close to the one that maximizes the vaccination coverage (green point) because we are assuming very similar rates of immunity loss for the vaccinated individuals of the three age groups.

However, increasing the differences between these rates results in greater distances between both points, as it can be observed in Supplementary Fig. S1 online, where the probabilities *p*_*i*_ are also different for each age group. In both figures, 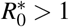 at the maximum coverage (green point) for the data sets from Belgium (1.0028, 1.0037) and Germany (1.0020, 1.0017), whereas 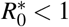 at this point for the data sets from Italy (0.99786, 0.99024), the Netherlands (0.99538, 0.98184), and Peru (0.99001, 0.97032). For the data set from Zimbabwe, 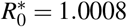 in Fig. 3 and 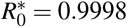 in Supplementary Fig. S1 online.

In Fig. 3 we can also see the vaccine allocations that result in the least disease containment (the worst strategies). In all the plots, these strategies result in values of 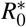 larger than 2 (orange region), while it is assumed to be equal to 2.5 at the DFE without vaccinated individuals. In Belgium and Germany, the orange region is at bottom of the contour plots which corresponds to a very low vaccination of the adults 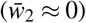. This age group makes up 57% of the population in both countries, and its total per capita contact rate is the highest in Belgium and very close to the highest in Germany. In Peru, the worst strategies (orange region) are clearly on left side of the contour plot, which corresponds to a very low vaccination rate of the age group [0, 18) 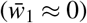, which makes up 36% of the population and has the highest per capita number of contacts. In Italy, the Netherlands, and Zimbabwe, the orange region is concentrated at the lower left corner, that is, the worst strategies correspond to allocate most of the available vaccines to the elderly 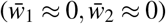. This age group either has the fewest contacts per capita (Italy, the Netherlands), or it is by far the least numerous group (Zimbabwe).

In Table 2, we can see that, as just explained above, the per capita vaccination rates 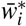 computed from (3) are all very close to 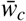 with 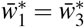 (because 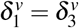). Moreover, these rates lead to the same vaccination coverage as the uniformly random vaccination with 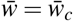 defining the (cf. Table 1). But, remarkably, they are clearly different from the vaccination rates 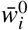 defining the minimum value of 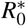 for the same value of 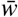 (blue points in Fig 3). The latter are the optimal per capita vaccination rates and they nicely correlate with the total contact rate of the age groups (see Supplementary Table S1 online). For instance, the variation in the magnitude of Zimbabwe’s vaccination rates is the opposite of that of the other countries, as is the order of the magnitude of its total per capita contact rates. Moreover, the similarity of the contact patterns of the data sets of Belgium and Germany, and those of the data sets of Italy and the Netherlands (see Methods), is reflected in the similarity of the values of the corresponding optimal per capita vaccination rates.

**Table 2.**
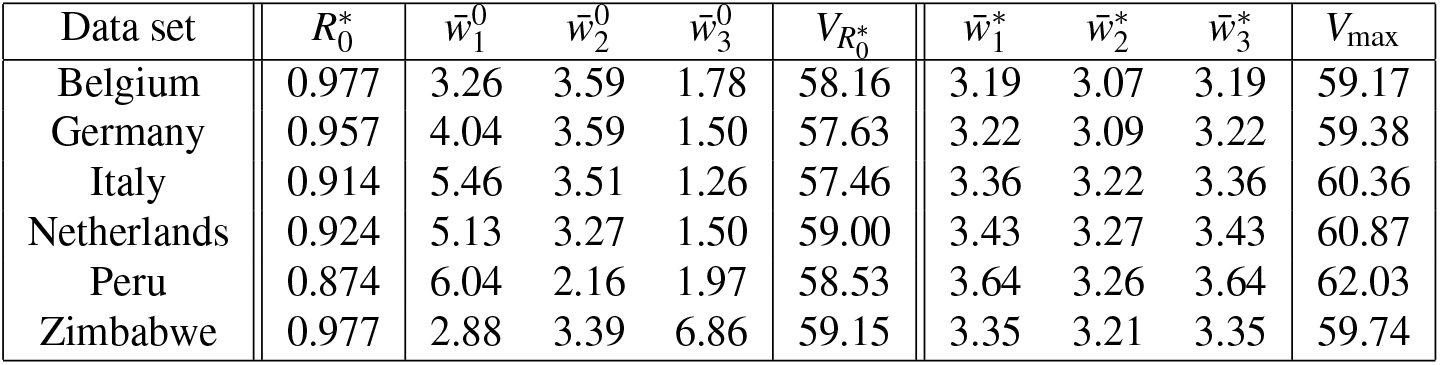
Mean vaccination coverage (in %) at the vaccination strategy 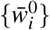 (in %) leading to the minimum 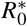 at the DFE, and at the vaccination strategy 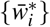 (in %) computed from Eq. (3) leading to the maximum mean vaccination coverage. In both cases, the mean per capita vaccination rate 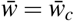, and 100% vaccine efficacy is assumed. Parameters: 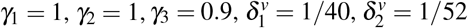, and 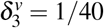. For each country, *β* is scaled such that *R*_0_ = 2.5 in the absence of vaccinated individuals.

In Fig. 3, we also observe that there is a distance between the blue point and the 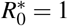 black curve, which is more evident in some of the six plots. This distance creates the opportunity for vaccination rates that can still guarantee a locally stable DFE but with a mean per capita vaccination rate below the critical rate obtained under the assumption of a uniformly random vaccination.

To illustrate this fact, in Fig. 4, we show contour plots for 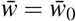 such that the rounded value of the minimum 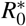 is equal to 0.996. As expected, in these figures this minimum (blue point) is very close to the 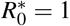 curve because, now, we are administering a lower number of vaccines. Considering Italy, for example, we obtain 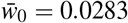, which, compared with 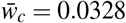, is equivalent to a reduction of the vaccination rate of 13.7%. Such a reduction of the vaccination rate can correspond to non-negligible savings. Peru is the country, among the ones considered, which gains the larger vaccination rate reduction, from 0.0354 to 0.0284, with a vaccination reduction of 19.8%. Table 3 summarizes the vaccination rates for 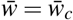 and 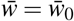 for the selected countries and the corresponding vaccination coverage. When we look for a vaccination strategy to bring the system to the disease-free equilibrium, taking into account the country’s contact patterns per age-group provides an opportunity to reduce the critical vaccination rate compared with the one needed considering homogeneous mixing. This also has a consequence on the estimation of the herd immunity coverage for each country. The consideration of contact patterns at the level of age groups, reducing the required vaccination rate, also reduces the level of vaccination coverage required, reducing in turn the herd immunity levels, as it is also shown in this table.

**Figure 4.**
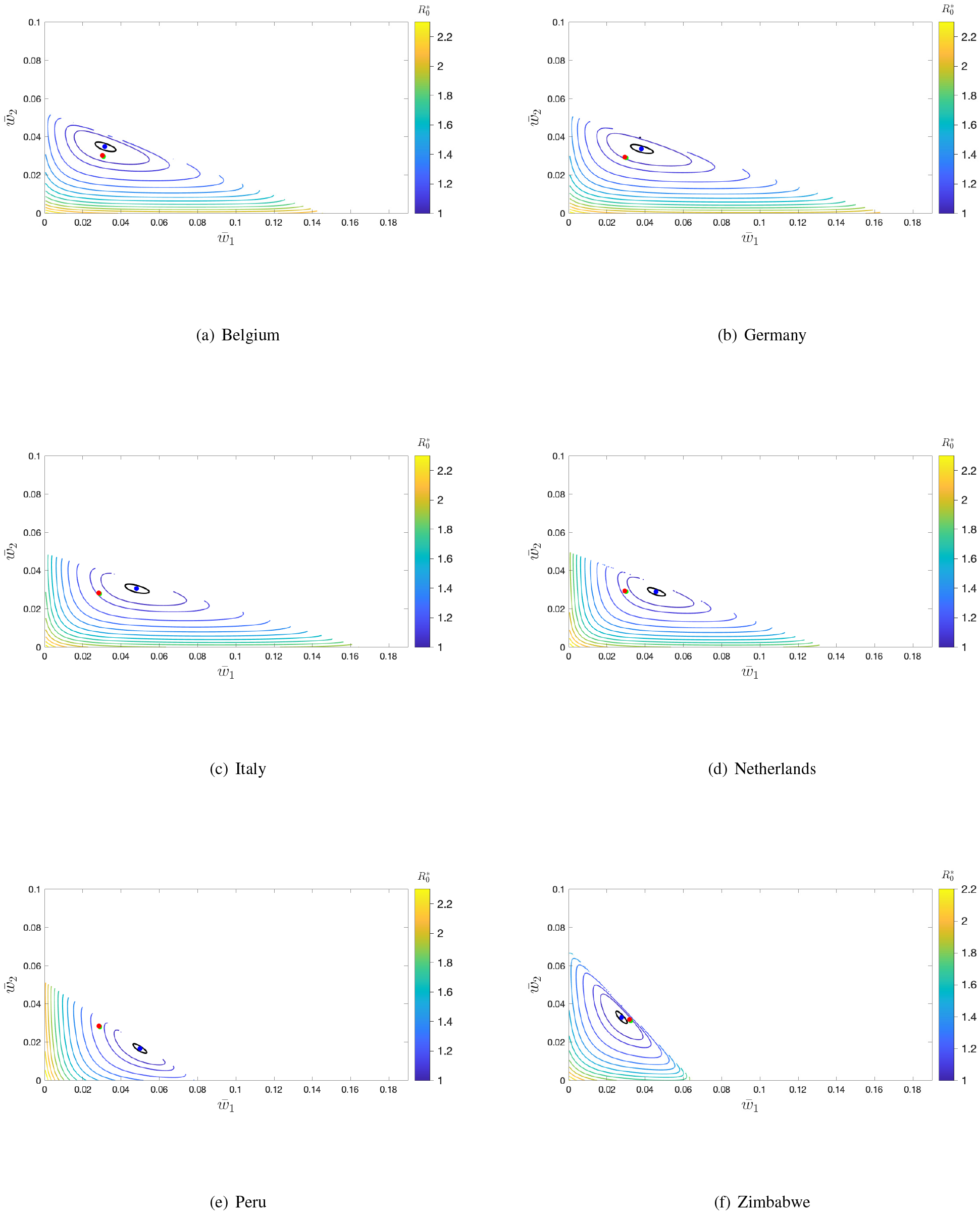
Contour levels of 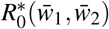 at the DFE of system (1) with *p*_*i*_ = 1 (*i* = 1, 2, 3) and for 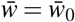, a value for which *R*_0_ ≈ 0.996 (see Table 3). Black level curve corresponds to 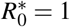. Blue point: Minimum of 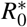. Red point: 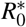 under uniformly random vaccination 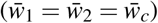. Green point: 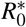 at the disease-free equilibrium with the maximum vaccination coverage. Parameters: 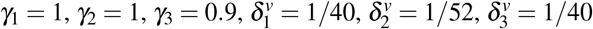. For each country, *β* is scaled such that *R*_0_ = 2.5 in the absence of vaccinated individuals.

**Table 3.** Mean vaccination coverage (in %) at the critical per capita vaccination rate 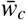 (in %) under the uniformly random vaccination strategy, and at the mean per capita vaccination rate 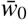 (in %) at the DFE for which 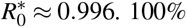 vaccine efficacy is assumed in both cases.

| Data set | $\bar{w}_c$ | $V_{\bar{w}_c}$ | $\bar{w}_0$ | $V_{\bar{w}_0}$ |
| --- | --- | --- | --- | --- |
| Belgium | 3.12 | 59.16 | 3.02 | 57.32 |
| Germany | 3.15 | 59.37 | 2.94 | 55.89 |
| Italy | 3.28 | 60.36 | 2.83 | 53.69 |
| Netherlands | 3.34 | 60.86 | 2.94 | 55.81 |
| Peru | 3.54 | 62.02 | 2.84 | 52.74 |
| Zimbabwe | 3.29 | 59.73 | 3.18 | 58.33 |

## Discussion and conclusions

During an ongoing epidemic like COVID-19, priorities are focused on immunizing in a short time as many people as possible, those working in front-line healthcare staff, in essential services, and those whose health conditions predispose to severe morbidity from infection. In contrast, in the long term, *R*_0_ can play a role in defining the minimum vaccination coverage for preventing new epidemic invasions^10,12^ by reaching the so-called herd immunity. It is currently not clear whether many countries could achieve such a herd immunity for COVID-19. Several reasons have been proposed for that: the limited availability of vaccines in many countries, the fact that immunity might not last forever, or the appearance of new variants of the virus that could change the herd-immunity threshold itself^31^. There is also a critical hesitation against vaccination arising from the spread of misinformation on the Internet^32^, which has been called COVID-19 infodemic^33^.

In this paper, we have assumed a limited supply of vaccines conferring waning immunity to deal with some of these issues. The aim is to see how herd immunity can be achieved in partially vaccinated populations whose individuals are classified in three age groups (youngsters, adults, and the elderly), when their contact patterns are taken into account. Moreover, by assuming similar disease rates for all age groups and countries, we have been able to assess the impact of social contact patterns on the critical vaccination coverage.

Such an impact has been analyzed by obtaining the set of per capita vaccination rates that minimizes 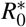, the basic reproduction number at the DFE with vaccinated individuals. Such a minimization is done under the assumptions of (1) a limited supply of vaccines given by the critical vaccination rate 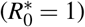 and (2) a uniformly random vaccination. Even though these are strong constraints, the first observation is that this minimizing set of rates defines a vaccination strategy that reduces 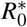 to values that are clearly below 1 (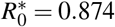 for the data set of Peru). So, our first conclusion is that there is room for an improvement in the vaccine distribution when demographic (population composition) and social aspects are considered. We estimate the vaccine reduction achieved following the 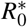-minimization strategy by decreasing even more the total availability of vaccines per unit of time in such a way that the minimum of *R*_0_ is close to 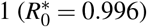.

A second conclusion is that, by adopting a vaccination strategy that minimizes 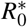, we are giving higher protection to those age groups that are more vulnerable to contract the infection in the absence of vaccination due to their social contact pattern. This vaccination strategy is in sharp contrast to the one that yields the maximum mean coverage of the population, which only depends on the rate of immunity loss and the probabilities of successful protection against infection. Precisely, age groups with the highest/lowest per capita vaccination rates at the minimum 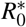 (see Table 2) correspond to those with the smallest/highest fractions of susceptible individuals at the endemic equilibrium without vaccination (cf. Fig. 2) which, in turn, correspond to the age groups with the highest/lowest total per capita contact rates. The data set from Zimbabwe is particularly interesting since it is the only data set where the highest total per capita vaccination rate corresponds to the 60+ years age group, while the fraction of population that it represents (4.4%) is the smallest one in all data sets. However, the per capita number of contacts of this age group is much higher than the one of the same age group in the rest of the data sets.

These findings are consistent with the well-known fact that vaccinating individuals with the highest numbers of contacts reduces the spread of an infectious disease. Indeed, minimizing 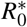 can be thought as a way to find effective risk-based allocations of a limited supply of vaccines. However, other criteria of optimality have been adopted in previous literature as, for instance, the one that chooses the vaccination strategy that allocates the fewest vaccines among all the strategies guaranteeing *R*_0_ *<* 1 when there is a sufficient supply of vaccines and permanent immunity^14^.

The results presented in the paper provide a first insight into the role of contact patterns in the spread of an infectious disease like COVID-19 which leads to a short-lived immunity, and in the optimal vaccination strategy based on the minimization of *R*_0_. The values of the disease parameters have been chosen to approximately mimic the COVID-19 infectious period (one week) and the current estimates of the possible duration of the immunity (about one year). For our study, we used a simple SIRV epidemic model that neglects relevant aspects of the COVID-19 dynamics, such as, for instance, the existence of a latent period and different levels of disease severity. Nevertheless, our analysis mainly focuses on the basic reproduction number for populations where only susceptible and vaccinated individuals are present. Therefore, the inclusion of more non-infectious compartments in the model will not change the paper’s main conclusions. Other modeling aspects such as the individual variation in susceptibility, or differences in social activity within age groups leading to different exposures to the virus, have also been neglected in the present study, even though they may contribute to an even larger reduction in the required vaccination coverage, as recent studies on disease-induced herd immunity against SARS-CoV-2 have revealed^16,34^.

## Data Availability

All data produced in the present study are available upon reasonable request to the authors

## Acknowledgments

The work of Caterina Scoglio has been supported by the National Science Foundation under Grant Award IIS-2027336. Joan Saldaña has been partially supported by Grant No. PID2019-104437GB-I00 of the Agencia Estatal de Investigación, Ministerio de Ciencia e Innovación of the Spanish government.

## Author contributions statement

J. Saldaña and C. Scoglio conceived the research, analysed the results, and wrote the manuscript. J. Saldaña also developed the formal analysis of the model and its numerical implementation.

