## Supplementary tables and figure for "Influence of heterogeneous age-group contact patterns on critical vaccination rates for herd immunity to SARS-CoV-2"

In Table S1, we report the contact data matrices used to derive our results. Contact data have been downloaded with the default settings, such as all contacts for day type, contact duration, contact intensity and all genders. Reciprocity, weigh by age, weigh by week/weekend, include supplemental professional contacts, and all locations have been checked.

In particular, reciprocity means that contacts are reciprocal and, hence, the total number of contacts per unit of time of individuals of age group $i$ with individuals of age group $j$ has to be equal to total the number of contacts per unit of time of individuals of age group $j$ with individuals of age group $i$. This is equivalent to impose that $c_{ij}f_i = c_{ji}f_j$ where $C = (c_{kl})$ is the contact matrix and $f_k$ is the fraction of the population in the age group $k$.

For each country, the population fraction $f_k$ of each age group used in our analysis is shown in Table S2.

| Country | Age group | contact $[0,18)$ | contact $[18,60)$ | contact 60+ | total contacts |
|---|---|---|---|---|---|
| Belgium | $[0,18)$ | 5.426 | 5.143 | 0.772 | 11.341 |
| | $[18,60)$ | 1.864 | 9.383 | 1.540 | 12.787 |
| | 60+ | 0.723 | 3.982 | 3.175 | 7.880 |
| Germany | $[0,18)$ | 4.045 | 3.907 | 0.567 | 8.519 |
| | $[18,60)$ | 1.219 | 6.216 | 0.998 | 8.433 |
| | 60+ | 0.408 | 2.299 | 2.127 | 4.834 |
| Italy | $[0,18)$ | 12.753 | 8.990 | 1.513 | 23.256 |
| | $[18,60)$ | 2.669 | 13.530 | 2.594 | 18.793 |
| | 60+ | 1.032 | 5.957 | 3.558 | 10.547 |
| Netherlands | $[0,18)$ | 11.181 | 5.649 | 0.681 | 17.511 |
| | $[18,60)$ | 2.108 | 10.714 | 1.491 | 14.313 |
| | 60+ | 0.775 | 4.548 | 3.464 | 8.787 |
| Peru | $[0,18)$ | 13.855 | 6.664 | 0.792 | 21.311 |
| | $[18,60)$ | 4.369 | 6.559 | 1.039 | 11.967 |
| | 60+ | 3.210 | 6.422 | 1.262 | 10.894 |
| Zimbabwe | $[0,18)$ | 5.580 | 3.811 | 0.524 | 9.915 |
| | $[18,60)$ | 4.025 | 6.617 | 0.825 | 11.467 |
| | 60+ | 5.861 | 8.737 | 2.007 | 16.605 |

**Table S1**. Daily contact matrices used in the numerical integration. The last column gives the sum of each row which is equal to the total per capita daily number of contacts.

| Age group | Belgium | Germany | Italy | Netherlands | Peru | Zimbabwe |
|---|---|---|---|---|---|---|
| $[0,18)$ | 0.207 | 0.179 | 0.172 | 0.219 | 0.361 | 0.491 |
| $[18,60)$ | 0.572 | 0.572 | 0.577 | 0.588 | 0.550 | 0.465 |
| 60+ | 0.221 | 0.249 | 0.251 | 0.193 | 0.089 | 0.044 |

**Table S2**. Population fraction of each age group for the considered countries.

### Additional contour plots

To complement the contour plots presented in the main text, Figure S1 shows a choice of values of the parameters that increases the heterogeneity among the three age groups: lower waning rates of immunity for the age groups 1 and 3, and slightly different probabilities of successful protection of the vaccine for each age group. Such heterogeneity implies that the vaccination strategy that maximizes the vaccination coverage under the constraint $\bar{w} = \bar{w}_c$ (green points) is now clearly distinguishable from the uniformly random vaccination strategy (red points).

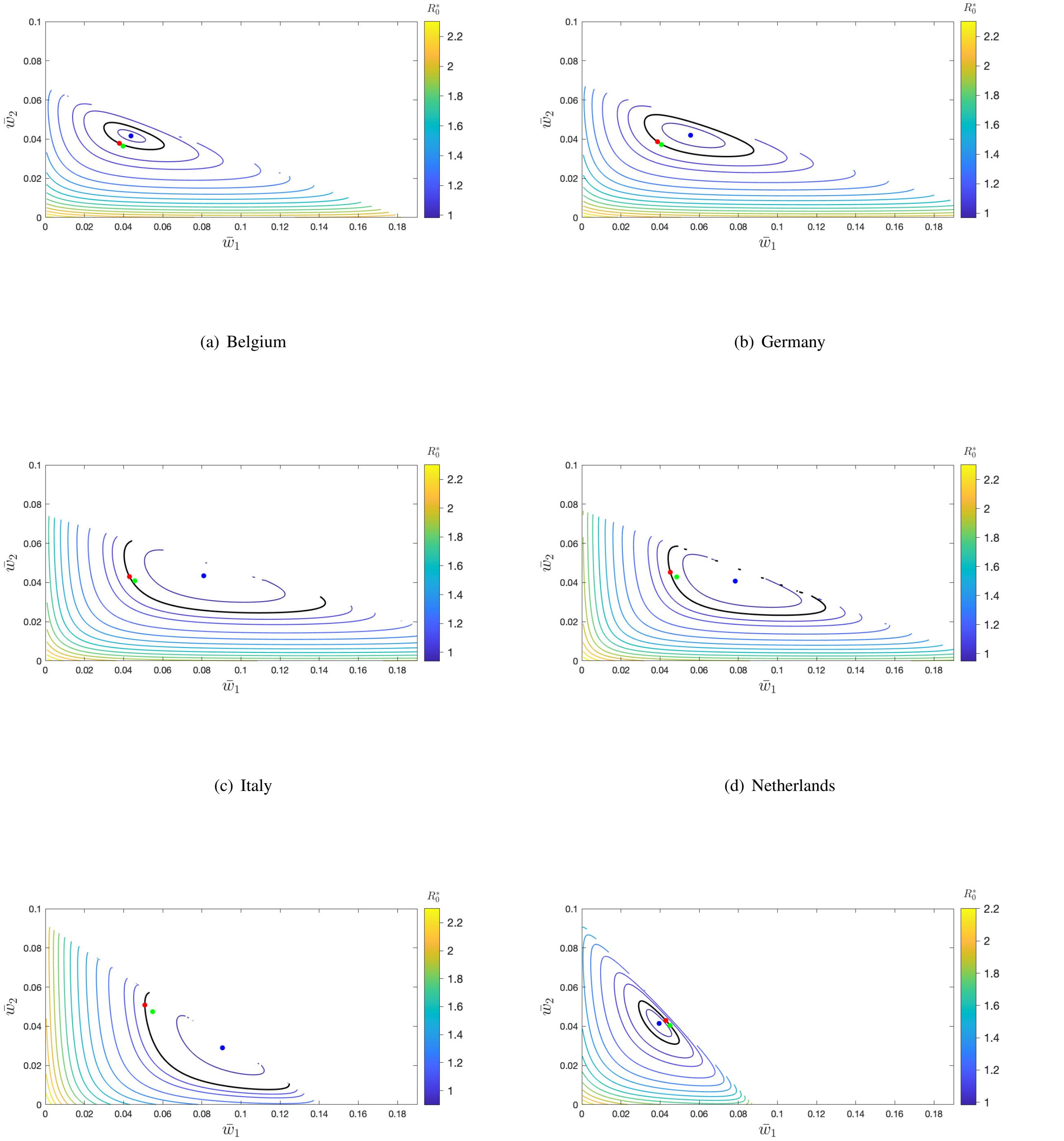


(a) Belgium

(b) Germany

(c) Italy

(d) Netherlands

(e) Peru

(f) Zimbabwe

**Figure S1**. Contour levels of $R_0^*(\bar{w}_1, \bar{w}_2)$ at the DFE of system (1) with $p_1 = 1$, $p_2 = 0.95$, and $p_3 = 0.9$. Black level curve corresponds to $R_0^* = 1$. Blue point: Minimum of $R_0^*$. Red point: $R_0^*$ under uniformly random vaccination ($\bar{w}_1 = \bar{w}_2 = \bar{w}_c$). Green point: $R_0^*$ at the disease-free equilibrium with the maximum vaccination coverage. Parameters: $\gamma_1 = 1$, $\gamma_2 = 1$, $\gamma_3 = 0.9$, $\delta_1^v = 1/26$, $\delta_2^v = 1/52$, $\delta_3^v = 1/26$. For each country, $\beta$ is scaled such that $R_0 = 2.5$ in the absence of vaccinated individuals.
